# Screening for anemia using multi-modal machine learning models on smartphones: protocol for a comparative accuracy study in rural India

**DOI:** 10.1101/2025.04.10.25325591

**Authors:** Shrey Desai, Kaushal Jesalpura, Sujay Kakarmath, Mayank Daswani, Sethuraman Venkatraman, Jim Taylor, Shruti Gadgil, Marc Wilson, Rajroshan Sawhney, Anaita Singh, Raghu Pullakhandam, Matthew Thompson

## Abstract

**Introduction:** Anemia, or low blood hemoglobin (Hb), affects one third of the world population, and is particularly prevalent in women and children in lower resource settings. However, screening for anemia is limited by the availability of accurate, easy to use, lower cost and non-invasive methods. We aim to generate research to support potential development of a point of care test to detect anemia using smartphone images of conjunctivae, tongue and nail beds as well as photoplethysmogram (PPG) signals, and comparing their accuracy to a standard laboratory assay, and, a point of care Hb assay.

**Methods & Analysis:** Cross-sectional comparative accuracy study of Adults and children (>1 year) presenting to hospital and outpatient care at SEWA Rural, a non-governmental organization providing healthcare to a rural, tribal population in Gujarat, India. Patients whose clinician has requested a complete blood count (CBC) will be recruited. Images of conjunctivae, tongue and nail beds, and a 30 second recording of fingertip PPG will be obtained using up to three different smartphones. A point of care Hb (HemoCue Hb 301) will be performed. Machine learning will be used to derive algorithms from images and/or PPG that predict anemia across a range of severities, and age groups. We will determine the mean absolute error and standard deviation of algorithms, compared to the laboratory value of Hb. We will also compare estimates of model accuracy using sensitivity, specificity and 95%CI compared to the reference test for different severities of anemia, and also to the point of care Hb assay. A total sample size of 2322 is estimated to provide a mean absolute error of < 1g/dL. Accuracy will be explored in subgroups including age, gender, pregnancy, presence of clinical features of anemia, and skin tone.

**Ethics and Dissemination:** The study was approved by SEWA Rural Institutional Ethics Committee on 31 March, 2023. The Health Ministry Screening Committee (HMSC) approved the conduct of this study. Findings from the study will be presented in the peer reviewed journal, and data will be made available for external researchers.

**Trial registration:** The study is registered with the Clinical Trial Registry of India (Registration number: CTRI/2023/07/055313).

**Strengths and Limitations:**

- Participants will include populations of high clinical importance, including children, pregnant women and those with moderate or severe anemia
- Smartphone data used to train models will include images from multiple body sites as well as photoplethysmogram (PPG data), in order to facilitate identifying the type and location of sensor data that provides most accuracy for hemoglobin estimation.
- Several smartphone models, with varying sensing capability and cost will be used to collect data, to understand potential impacts of smartphone sensor quality/signal processing on Hb estimation.
- Non-consecutive recruitment may overestimate estimates of diagnostic test performance that differ from when a test is used in consecutive sampling
- Study participants may differ from those in other parts of India and other countries in characteristics such as skin tone, cause(s) of anemia, appearance of hands or eyes which may limit generalizability of findings.

## Introduction

Anemia, a reduced level of hemoglobin (Hb), affects between a quarter and a third of the world’s population and contributes to increased morbidity and mortality, decreased work productivity, and impaired neurological development (1, 2). Women of reproductive age (15-49 years) are at a particularly high risk of anemia compared to men, and maternal anemia can lead to adverse maternal and perinatal outcomes (3). Almost 300 million children younger than 5 years worldwide are anemic, with most living in low and middle-income countries (LMICs) (4). The most common causes of anemia worldwide are iron deficiency, deficiencies in other hematinic nutrients such as vitamin B12 and folic acid, infections, hemoglobinopathies, and malaria (2). Screening and diagnosis of anemia, together with clinical history, can lead to targeted interventions to treat causes of anemia (such as treatment of parasitic infections), use of iron supplements and/or blood transfusions, and to identify need for higher levels of medical care. The World Health Organization’s (WHO) Global Nutrition Target has a goal of reducing anemia by half by 2025 (4). While current efforts have led to improvements in reducing anemia prevalence, only three out of 82 LMICs are on target to meet these goals (3).

India faces particularly high rates of anemia. The recent National Family Health Survey for 2019-21 found that anemia was present in 57% of women of reproductive age, and 67% of children under 5 years of age (*6*). Rates of anemia are even higher among tribal women and children, with prevalences ranging from 78 to 96% (7). The Government of India started the multifaceted Anemia Mukt Bharat program in 2018 to reduce the burden of anemia, targeting children, adolescents and women of reproductive age group; one of the program’s key interventions is screening for anemia in community settings using digital and other point of care diagnostic methods (8).

Clinical examination of the tongue, eye, or fingernails for pallor has very poor sensitivity although moderate specificity for detecting anemia (9). Therefore anemia is typically diagnosed using a blood test. The gold standard method for anemia diagnosis is measurement of venous blood Hb using automated hematology analyzers. However, several tests have been developed as alternatives to formal laboratory testing for use in community settings, ranging from the simple WHO hemoglobin color scale, to various point of care assays utilizing fingerstick tests and portable analyser devices. These show varying accuracy, and are also limited by the need for blood sampling, costs of devices and disposable items (10, 11, 12). In addition, hemoglobin measurements in capillary blood using POC devices reported produce erroneous estimates resulting in large variations in anemia prevalence (13, 14, 15).

Recently, several non-invasive methods have been developed for detecting anemia without the need for blood sampling (11, 16). These methods rely on detecting anemia from spectrophotometry or photoplethysmography (PPG) (17). With the increasing use of smartphones by healthcare workers in lower resource settings, several research studies have explored whether the cameras of smartphones could be used to detect anemia. Indeed, some researchers have used the smartphone camera to detect PPG signals by measuring the reflectance or transmittance of LED light signals from blood in the fingernail bed (16, 19). Others have explored using software algorithms to detect pallor (indicating reduced Hb) from camera images of parts of the body (20, 21, 22). While accuracy of some of the smartphone based methods have been promising, none have progressed to more rigorous evaluation or regulatory approval, and none are in widespread use.

This protocol describes a study which has the primary objective of developing machine learning algorithms that can potentially be used to predict Hb concentration from smartphone images (of conjunctivae, tongue and nail beds) and PPG signals, as compared to venous Hb measured by automatic hematology analyzer and a point of care Hb measurement device.

## Material and analysis

### Study design

This prospective study will collect data to train and tune machine learning algorithms to detect anemia using smartphone images and PPG signals from adults and children presenting to outpatient and inpatient clinical settings in whom a CBC has been requested as part of their routine health care. The accuracy of these models will be compared to reference test of Hb measured by laboratory analyser across different levels of anemia (mild, moderate and severe anemia). We will also examine accuracy across subgroups of age, gender, pregnancy status and skin tone. We will also compare the accuracy of machine learning algorithms for detecting anemia, with that of clinical history and examination findings, and a commonly used point of care Hb measurement device.

### Patient and Public Involvement

Patients were not involved in designing or conducting this study.

### Study setting

The study will be conducted at SEWA Rural (Society for Education, Welfare and Action–Rural), a voluntary development organization involved in health and development activities in rural tribal areas in the Jhagadia block in southern Gujarat. SEWA Rural supports the overall development of the rural tribal population residing in Bharuch District (population: 250,000). The SEWA Rural hospital delivers outpatient and inpatient secondary level health services for pediatrics, maternal health, ophthalmology, general medicine and general surgery. Rural and tribal patients from more than 2,500 surrounding villages use the health services offered by SEWA Rural either free or at highly subsidized rates. The laboratory at the main hospital provides comprehensive services in hematology, clinical chemistry and serology along with essential services in cytopathology and blood banking.

### Participants

Potential participants will be identified from patients (age 1 to 89 years of age) who have a complete blood count (CBC) ordered by a clinician during routine outpatient and inpatient clinical care. This will include patients with a broad range of potential indications for CBC, including those with and without symptoms, and from a variety of medical, surgical, pediatric, maternal health areas of the inpatient and outpatient facilities at SEWA Rural.

Once CBC results are available, a list of potentially eligible participants will be generated on a daily basis by study investigators, and categorized by age and Hb level. A study Investigator from SEWA Rural will share information about the study in English, Gujarati, or Hindi with potential participants. For those interested in participating in the study, the study investigator will verify that the participant is fully informed, and has signed the consent form. Potential participants will be selected from three age groups (1-17, 18-49, 50-89 years) and four categories of Hb level (normal, and mild, moderate, and severe anemia) based on the CBC result. This form of recruitment was selected over consecutive recruitment in order to ensure that sufficient participants are recruited for all age and Hb categories. Participants will not receive monetary compensation for participation in this study.

### Inclusion and exclusion criteria

Participants must meet all the following criteria to be eligible for participation:

1. Age 1-89 years who are getting CBC as part of routine care as advised by treating physician at SEWA Rural
2. Able to speak and read English, Gujarati, and/or Hindi or has a legal guardian who can speak and read English Gujarati, and/or Hindi
3. Willing and able to provide informed consent in English, Gujarati, and/or Hindi or has a legal guardian (or parent) who can provide informed consent in English, Gujaratis, and/or Hindi
4. Willing to comply with evaluations and protocols

Participants with any of the following criteria will be excluded from participation:

1. Have any additional condition or situation that the Investigator or designee determines as inappropriate for participation in this study.
2. Amputation or severe damage to all fingers of hand where data will be collected.
3. Unwilling or unable to remove nail polish/art/coverings or anything else that obscures the nails.
4. Unwilling or unable to provide a blood sample for CBC.

### Reference test

Measurement of venous blood Hb derived from cyanide free SLS method from Sysmex XN-L series cell counter (Sysmex XN-550, Sysmex Corporation, Kobe, Japan) will be used as the reference test. The SEWA Rural laboratory has implemented a rigorous internal quality control system for the cell counter by utilizing a three-level control strategy to ensure calibration at three levels of Hb (5.5, 12, and 16.5 g/dL) and coefficient of variation (CV) of the assay < 1%), which will ensure the quality on all days prior to the sample analysis throughout the study period. The Sysmex cell counter also provides the following in addition to hemoglobin, namely hematocrit (Hct), red blood cell count, mean corpuscular volume (MCV), mean corpuscular hemoglobin (MCH), mean corpuscular hemoglobin concentration (MCHC), and red cell distribution of width (RDW). The presence of hemoglobinopathies will be noted from patient history and/or previous reports noted in the laboratory records. For the purposes of data collection, the following cutoffs will be used to categorize Hb levels in terms of presence and severity of anemia: Norma, Hb >=12.0 g/dL; mild anemia Hb between 10.0 - 11.9 g/dL; moderate anemia Hb between 7.0 - 9.9 g/dL; severe anemia Hb < 7.0 g/dL. During analysis however, we will examine performance based on the WHO cutoffs and categorizations of anemia which take into account age, gender and pregnancy status (23).

### Study Procedures

After the consent form has been signed, the study investigator will obtain all other information using a specially designed data collection mobile phone application (Figure). Data collection from participants will be conducted at a separate location inside the laboratory, which is an interior room. Ambient lighting will be recorded using a standard luminometer (TESTO 540 Pocket sized Luxmeter, Testo, West Chester, USA), daily during each data collection period.

The study investigator will obtain demographics and medical history (including age, gender, religion, occupation), location and reason for health care encounter, presence of pregnancy, smoking history, and presence of symptoms potentially associated with anemia (Table 1). This will involve self report, with no review of participant medical records. Next, the study investigator will conduct a physical examination, including measurement of height and weight (using standard clinical grade equipment), temperature (Medtech TMP11 Portable Digital Thermometer, Medtech Life Pvt Ltd, Mumbai, India), heart rate and O2 saturation (OXYGARD, Pulse Oximeter OG 05, Medtech Life Pvt Ltd, Mumbai, India)), and in adults only systolic and diastolic blood pressure using an automatic arm cuff sized appropriately to the arm of the participant (Novacheck BP-18, Medtech Life Pvt Ltd, Mumbai, India)) (Table 3). Skin tone will be measured on the forehead using a spectrophotometer (CAPSURE, Pantone RM200QC Portable Spectrocolorimeter, Panton, Carlstadt, NJ, USA). The study investigator will examine the participant’s eyes, mouth, fingers for signs of anemia, nail abnormalities and other features.

**Table 1:** Data collected on demographics, medical history, and symptoms.

|  |  |  |
| --- | --- | --- |
| **Demographics** | **Age** | years/months |
|  | **Sex (at birth)** | Female<br>Male<br>other |
|  | **Caste** | Scheduled tribe (tribal-indigenous)<br>Scheduled caste<br>Other backward class<br>General |
|  | **Religion** | Hindu<br>Muslim<br>Christian<br>other |
|  | **Occupation** | Farming<br>Job<br>Work in other occupation<br>Housewife<br>Studying<br>Unemployed/Out of school<br>other |
| **Details of current health care visit** | **Setting** | Outpatient<br>Inpatient |
|  | **Reason for attending / admission** |  |
|  | **Service attended** | Pediatrics<br>OBGYN<br>Internal Medicine<br>General Medicine<br>General surgery<br>Ophthalmology<br>other service |
| **Medical history** | **Pregnancy status (females only)** | ☐ Currently pregnant<br>Gestational age: _____ |
|  | **Smoking and alcohol use** | Smoking history<br>☐ Currently smoking<br>☐ Previously a smoker<br>☐ Never smoked<br>Alcohol intake<br>☐ Currently consuming<br>☐ Previously consumed<br>☐ Never consumed |
|  | **Past Medical History** | Hypertension<br>Diabetes Mellitus type 1 or 2<br>Preeclampsia<br>Anemia<br>Disorders of blood (sickle cell, thalassemias) IF Yes, type of blood disorder: _____<br>Cancer or malignancy<br>other |
| **Current symptoms** | **Adults** | Tired or lack of energy<br>Shortness of breath at rest or on exertion<br>Fast heart beat, particularly during exercise<br>Losing weight<br>Chest Pain<br>Lightheaded or Dizziness<br>Headaches<br>Leg cramps<br>Brittle hair or nails<br>other |
|  | **Children (<18 yr)** | Looks pale<br>Losing weight or not gaining weight<br>More tired or sleepy than usual<br>Out of breath or difficulty breathing<br>other |

**Table 2:** Estimated sample sizes: Number of participants needed to have a power of 0.9 to detect a MAE in Hb measurement that is significantly < 1.0 g/dL, with multiple postulated MAE and standard deviations of the error (one-sided P value < 0.025)

| **SD** | **$\mu_{error} =$**<br>**0.1 g/dL** | **$\mu_{error} =$**<br>**0.15 g/dL** | **$\mu_{error} =$**<br>**0.20 g/dL** | **$\mu_{error} =$**<br>**0.25 g/dL** | **$\mu_{error} =$**<br>**0.50 g/dL** | **$\mu_{error} =$**<br>**0.65 g/dL** |
| --- | --- | --- | --- | --- | --- | --- |
| **SD=1.5**<br>**g/dL** | 32 | 35 | 39 | 44 | 97 | 195 |
| **SD=2.0**<br>**g/dL** | 54 | 61 | 68 | 77 | 171 | 346 |
| **SD=2.5**<br>**g/dL** | 84 | 93 | 105 | 119 | 265 | 539 |
| **SD=3.0**<br>**g/dL** | 119 | 133 | 150 | 171 | 381 | 774 |

\* AI = AI reading; Rf = Reference (ground truth) reading; $\mu_{error}$ = MAE; SD = standard deviation of AI and ground truth readings.

**Table 3:** Data collected on research location, vital signs, and examination findings.

|  |  |  |
| --- | --- | --- |
| **Details of data collection location** | **Place of data collection** | Indoor<br>Outdoor<br>other |
|  | **Location of data collection** | Laboratory<br>Ward<br>Outpatient department<br>other |
|  | **Time of data collection:** | 6 AM to 12 PM<br>12 PM to 6 PM<br>6 PM to 12 AM<br>12 AM to 6 AM |
|  | **Season of data collection** | Summer<br>Monsoon<br>Winter |
|  | **Light intensity** |  |
|  | **Ambient temperature (°C)** |  |
| **Vital signs** | **Height/length (cm)** |  |
|  | **Weight (kg)** |  |
|  | **Blood pressure (>=18 yr)** | Systolic_____mmHg<br>Diastolic_____mmHg |
|  | **Skin tone (individual typology angle, ITA)** |  |
|  | **Body temperature (°C)** |  |
| **Physical Examination findings** | **Nails** | Nail polish, henna or any other color applied<br>Koilonychia<br>Clubbing<br>Fungal infection<br>Dirt under nail<br>Long nails<br>Nail cap applied<br>Unkept nails<br>Nails are chewed |
|  |  | Brittle or spoon-shaped nails<br>Other |
|  | **Fingers/hands** | Cuts<br>Dirt<br>Worn out finger creases<br>Discolouration<br>Henna<br>other |
|  | **Conjunctivae/eyes** | Pallor<br>Redness<br>Kajal applied<br>Contact lens<br>Red eye (inflammation of conjunctivae)<br>Other |
|  | **Tongue** | Pale tongue<br>Soreness of tongue<br>Cracks at corner of lips<br>Tobacco stains<br>other |
| **Blood test results** | **Point of care result** | Hemoglobin _____g/dL |
| | **Laboratory complete blood count** | White blood cells _____$10^3/\mu\text{L}$<br>Red blood cells _____$10^6/\mu\text{L}$<br>Hemoglobin _____g/dL<br>Hematocrit _____%<br>MCV _____fL<br>Mean corpuscular Hb _____pg<br>Mean corpuscular Hb concentration ____g/dL<br>Platelets _____$10^3/\mu\text{L}$<br>Red cell distribution of width _____% |
|  | **Presence of sickle cell trait/disease** | Disease<br>Trait<br>Negative<br>Not known |

The study investigator will then use one of three smartphones provided specifically for the research study to obtain images of conjunctivae (from one eye), tongue, fingernails (from one hand), and a 30 second video of PPG from the index finger. For participants who have a color alternation of the index finger that they are unable to remove (e.g henna, nail polish, tattoos), another finger on the same hand, without color alternation, will be used. The three smartphones selected for data collection in this study, namely Samsung A14, Samsung A05 & Samsung S24 (Samsung Electronics Co Ltd, Suwon, S Korea) were selected based on the range of cost and image quality etc that they provide. The lower cost models (A14, A05) are widely used in India, whereas the higher cost model (S24) has technical advantages (e.g. higher dynamic range for video through HDR10+ being supported).

Finally, the research staff will measure participants’ Hb with the HemoCue Hb 301 System (HemoCue AB, Angelholm, Sweden) point of care anemia test using blood remaining from the venous sample provided for their CBC. In cases where insufficient venous blood is available, no additional samples will be collected. The results of the Hemocue test will be entered in the data collection app, but will not be shared with the participant or clinical staff.

No review of clinical records will be performed other than obtaining results of the CBC from the laboratory. The research staff performing the data collection, including obtaining smartphone images, clinical examination, and point of care test operation, will not be blinded to results of the laboratory CBC result, nor clinical reason for obtaining the CBC, which will be available prior to study procedures commencing. Research staff and laboratory staff will not have access to any results of Hb prediction from machine learning models.

### Data upload and storage

A copy of the de-identified data will be securely transferred to Google for algorithm development and analysis.

### Algorithm development

In order to train our machine learning algorithms, we will first partition the data into training (50%), and testing (50%) sets all of which will be stratified for Hb levels, age, gender, and pregnancy status. The testing set will remain held-out, and all model development will be performed on the training set, some subset of which will be used for model validation. We do not anticipate that the training partition will be sufficient to train a large model de novo for the task. Therefore, we will focus on fine-tuning existing vision models trained for other general tasks, and leverage existing foundational vision embedding models such as Contrastive Captioner (CoCa) (24). We will also compare against baselines that extract red pixel values from specific parts of the image (e.g. selecting the best bounding boxes on images from fingernails, tongue or conjunctivae) and training regression models using those (18).

### Statistical analysis plan

We will examine performance of algorithms within each subset of normal Hb, and anemia (mild, moderate, severe) using standard metrics such as R^2^, intra class coefficient (ICC), Bland Altman plots, and Pearson’s correlation coefficient (25). We will then measure mean absolute error (MAE) and its standard deviation (SD), as well as 95% confidence interval (95%CI) of estimated Hb. The 95% confidence interval will be computed by bootstrapping, resampling 1,000 times with replacement. A one sample t-test will be used to determine if the MAE is less than 1.0 g/dL (one-sided P < 0.025). If the distribution of the data are skewed, a Wilcoxon signed rank test will be used to determine if the MAE is < 1.0 g/dL.

To assess the clinical accuracy of our model, the sensitivity and specificity and associated 95% confidence intervals will be determined in various subgroups, defined by age, gender and pregnancy status and with different thresholds for defining mild, moderate and severe anemia. The different subgroups and definitions of anemia status will be based on WHO guidelines but modified where necessary based on data availability (23). Furthermore, accuracy of our model will also be compared with that of two other index tests, namely the presence of anemia noted on clinical features, and Hb measured using the HemoCue point of care device. For all three of these index tests (ie model, clinical features, and HemoCue), the comparison will be to the reference test Hb level. Accuracy will be compared among each of the above age, anemia, gender and pregnancy groups (Table 1) including sensitivity and specificity (and 95%CI). We will also explore accuracy of algorithms (compared to the reference test Hb) for the following subgroups: i) Presence of symptoms and physical signs, ii) skin tone, and iii) type of anemia defined as microcytic (MCV < 80 fL), normocytic (MCV 80-100 fL), macrocytic (MCV > 100 fL), and/or Sickle cell anemia.

### Sample size

The study sample size is largely driven by the large amount of data needed to train machine learning algorithms using data collected by each methodology to predict Hb levels. Models that are developed and tuned will then be validated in a test set that includes data on participants that were not used for training or tuning the algorithms. The test set will include enough participants in each of 3 age groups to have sufficient power to detect a mean absolute error (MAE) that is statistically significantly < 1 g/dL.

In determining the needed sample size for each of the 3 age groups in the test set, we do not have current estimates of MAE or SD available from existing image recognition or PPG software models which could be used to inform sample size calculations. The estimated sample sizes needed to identify a MAE <1 g/dL of estimated Hb to ground truth Hb, across a range of errors and standard deviations is displayed in Table 2. Given 3 age groups of primary interest (1-17, 18-49, 50-89 years) and range of levels of hemoglobin/severity of anemia and based on the highest MAE and SD, and including participants needed for algorithm training and tuning, the overall minimum study sample size is 2322 (774 per age group x 3 age groups = 2,322 in the test set). We intend to enroll a study sample that includes approximately 25% of participants with no anemia, 25% with mild, 25% with moderate, and 25% with severe anemia. However, enrollment to age groups and Hb levels will depend on patients attending the SEWA Rural hospital and outpatient clinics, and may not precisely match these approximate targets.

After recruiting approximately 500 participants we will i) Build preliminary models to determine the correlation, standard deviation, mean error of our models, ii) Evaluate the distribution of participant characteristics to the primary groups of interest including age-group, pregnancy trimester, gender and skin tone. At that time, target total enrollment will be adjusted if needed, to ensure a 90% power to detect a MAE of < 1 g/dL. During interim analyses, we will perform further power calculations for each of our subgroups of interest (e.g., age and anemia severity) and evaluate if we are powered to test against the model-specific hypotheses:

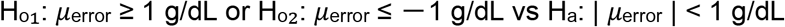

If we determine our subgroup performance has sufficient power with fewer than 774 participants per group, the study protocol will be modified to accommodate such changes.

## Ethics and Dissemination

The study has been approved by SEWA Rural’s Institutional Ethics Committee on 31 March 2023. The study was registered with the Clinical Trial Registry of India (Registration number: CTRI/2023/07/055313). The Health Ministry Screening Committee (HMSC) approved the conduct of this study. Findings from this research will be presented at appropriate conferences and will be submitted for publication in peer reviewed journals.

## Data Availability

All data produced in the present study are available upon reasonable request to the authors

## Authors’ contributions

SD, KJ, MD, SK,, RP, SG, JT, AS and MT conceived the study and SD, KJ, MD, SK and AS were involved in acquiring funding. The study methodology was conceived by SD, KJ, MD, SK, RP, SG, JT, AS and MT. Authors SD, KJ, MD, SK, SV, RP and SG obtained resources and MD, SV, RS and SG provided software and/or had input into the app development needed for the study. MD and JT conducted analyses for sample size calculation. SD, KJ, MD, SK, SV, SG and MT and MW will be conducting the investigation and SD and KJ are supervising on-site data collection study activities. MD, JT, MW and MT will be responsible for data analyses. SD, KJ, MD, SK, JT and MT wrote the original draft of the manuscript, and all authors reviewed and edited the final draft of the manuscript.

## Funding statement

This work was supported by the Bill & Melinda Gates Foundation, Seattle, WA to SEWA Rural [INV-043120:Inclusive and Representative R&D Space w for Digital Health Tools]. Google LLC did not provide any direct funding for this study, but provided in-kind support including clinical research expertise, engineering expertise, and analyses

## Competing interest statement

SD, KJ are employees of SEWA Rural and have received grants from the Bill and Melinda Gates Foundation for this study.SK, MD, JT, SD and MT are all employees of Google, and all have shares in Alphabet. SV - is an employee of Argusoft India Limited. AS and RP have nothing to declare

---

**Figure: Examples of screenshots from data collection app**

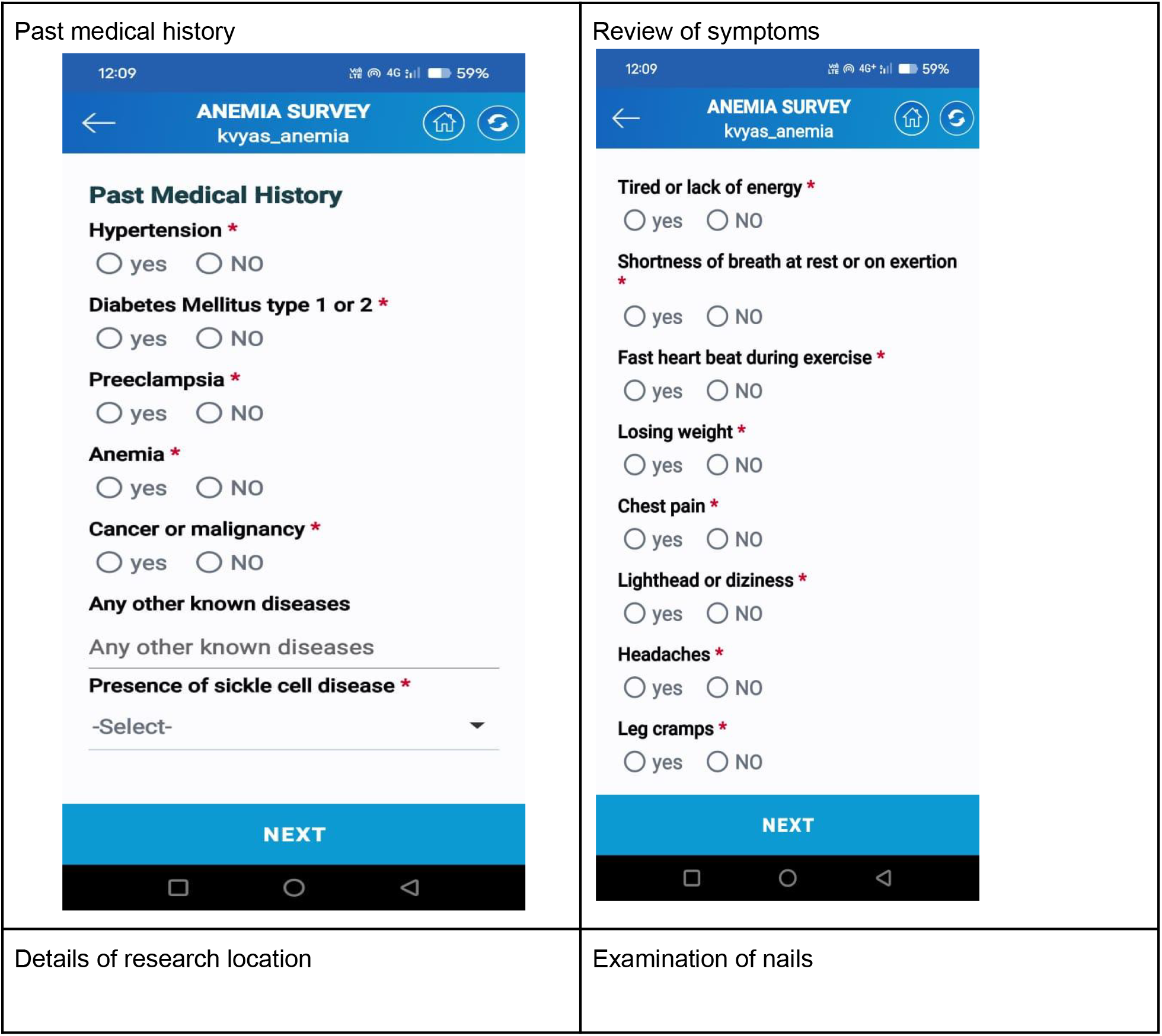
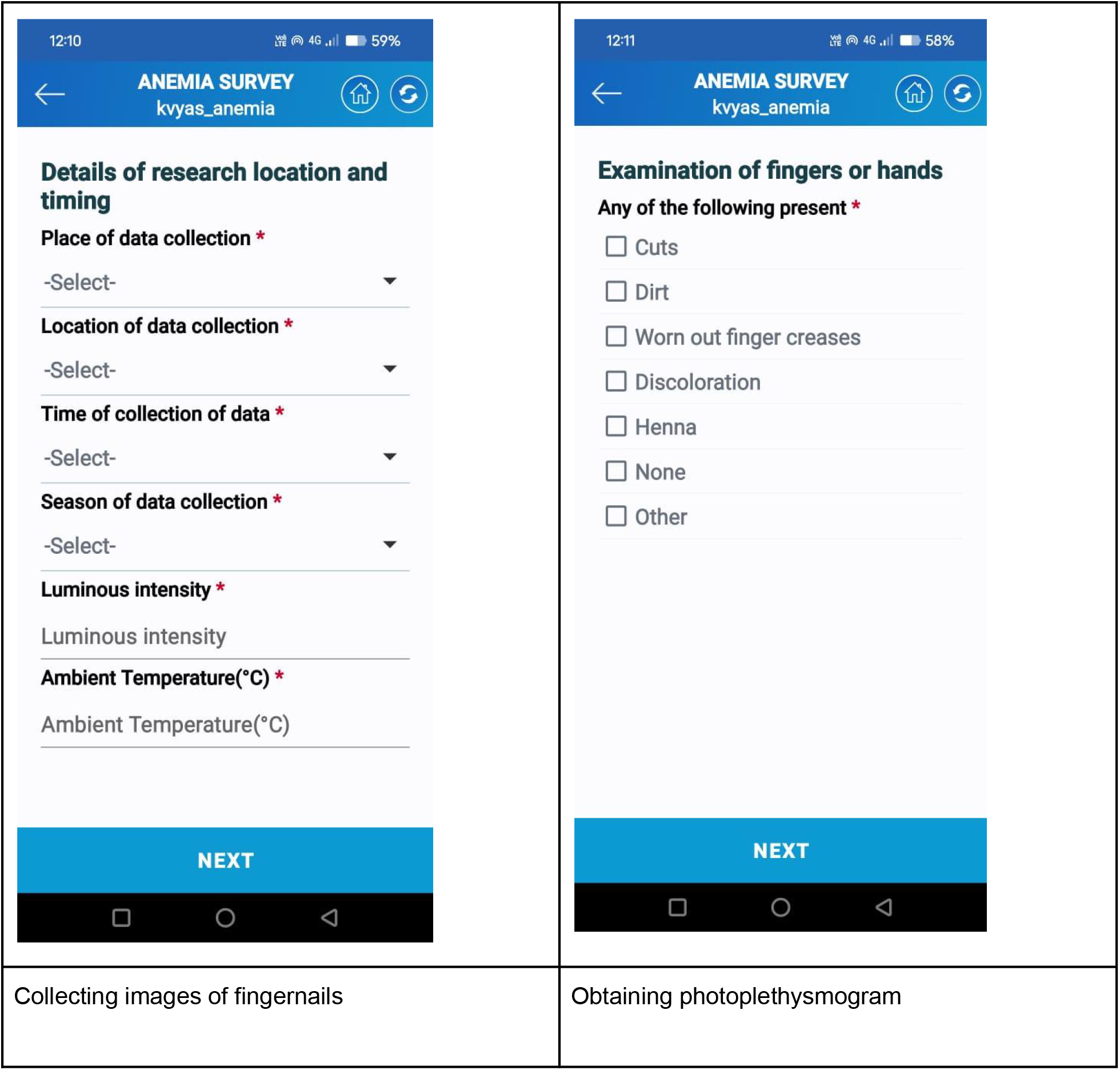
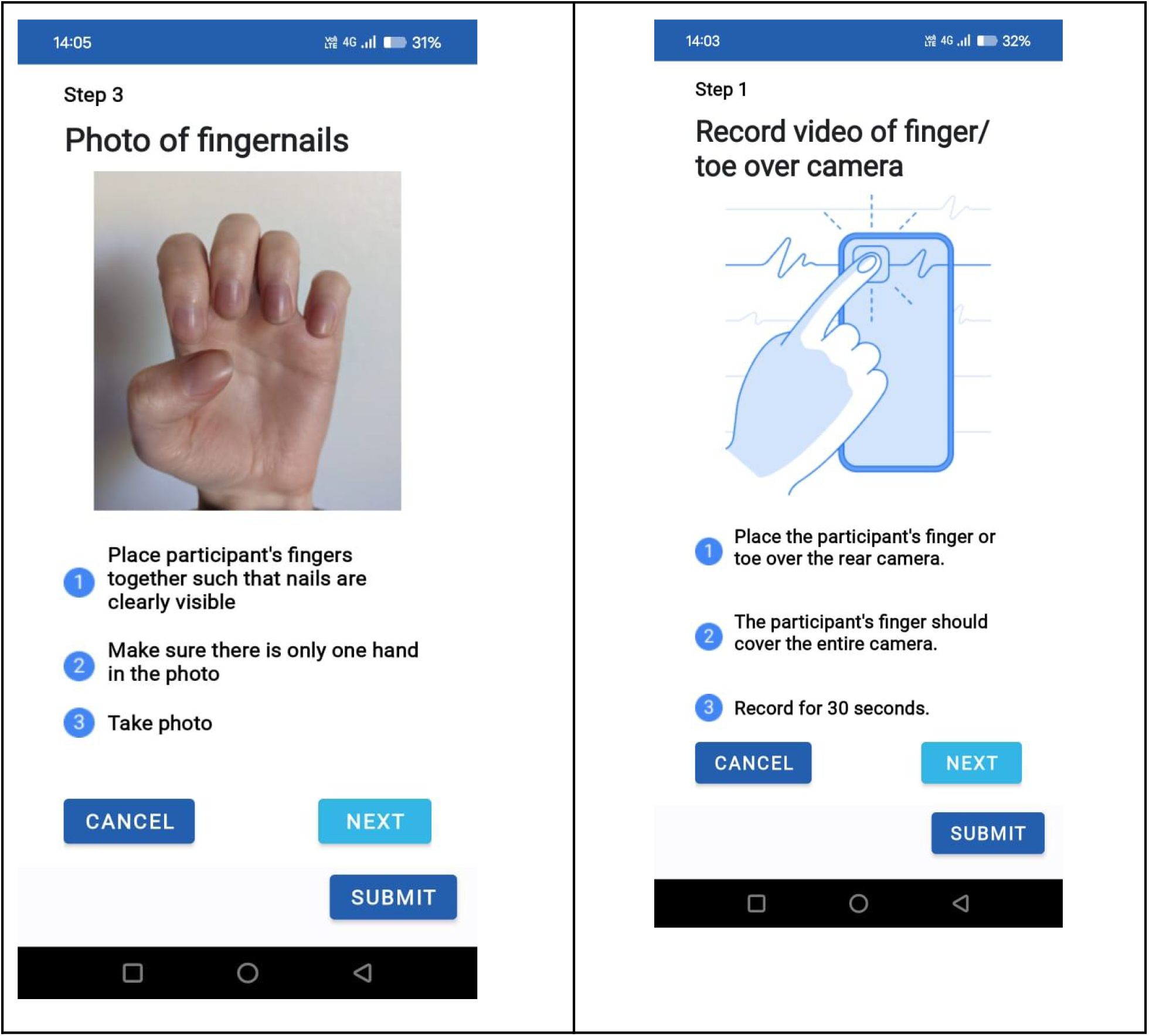

## Notes

### Author Declarations

The study has been approved by SEWA Rural Institutional Ethics Committee on 31 March 2023. The study was registered with the Clinical Trial Registry of India (Registration number: CTRI/2023/07/055313). The Health Ministry Screening Committee (HMSC) approved the conduct of this study

